# Leveraging the Genetics of Psychiatric Disorders to Prioritize Potential Drug Targets and Compounds

**DOI:** 10.1101/2024.09.24.24314069

**Authors:** Nadine Parker, Elise Koch, Alexey A. Shadrin, Julian Fuhrer, Guy F. L. Hindley, Sara E. Stinson, Piotr Jaholkowski, Markos Tesfaye, Anders M. Dale, Thomas S. Wingo, Aliza P. Wingo, Oleksandr Frei, Kevin S. O’Connell, Olav B. Smeland, Ole A. Andreassen

## Abstract

Genetics can inform biologically relevant drug development and repurposing which may improve patient care. Here, we leverage the genetics of psychiatric disorders to prioritize potential drug targets and compounds. We used the genome-wide association studies of four psychiatric disorders [attention deficit hyperactivity disorder (ADHD), bipolar disorder, depression, and schizophrenia] and genes encoding drug targets. We conducted drug enrichment analyses incorporating the novel and biologically specific GSA-MiXeR tool. We conducted multiple molecular trait analyses using large-scale transcriptomic and proteomic datasets sampled from brain and blood tissue. This included the novel use of the UK Biobank proteomic data for a proteome-wide association study of psychiatric disorders. With the accumulated evidence, we prioritize potential drug targets and compounds for each disorder. We reveal candidate drug targets associated with a single or multiple disorders and that implicate glutamate signalling. Drug prioritization indicated genetic support for psychotropic medications including several top ranked antipsychotics for schizophrenia. We also observed genetic support for commonly used psychotropics for psychiatric treatment (e.g., clozapine, duloxetine, and lithium). Revealed opportunities for drug repurposing included cholinergic drugs for ADHD, estrogen modulators for depression, and matrix metalloproteinases for ADHD and depression. Our findings indicate the genetic liability to schizophrenia is associated with reduced brain and blood expression of *CYP2D6,* a gene encoding a metabolizer of drugs and neurotransmitters, suggesting a genetic risk for poor drug response and altered neurotransmission. Our extensive analyses highlight the utility of genetics for informing drug development and repurposing for psychiatric disorders providing novel opportunities for improving patient outcomes.

**Graphical Abstract:** Depicted is the series of analyses conducted to generate a list of prioritized drug targets and compounds. First pairings of genome-wide association study (GWAS) traits with drugs are generated using enrichment analyses. Next a series of molecular trait analyses are conducted to generate and rank list of potential drug targets for each GWAS trait. Finally, enrichment and molecular trait results are combined to generate a ranked list of prioritized drugs for each GWAS trait based on supporting genetic evidence. ADHD = Attention deficit hyperactivity disorder, BIP = Bipolar disorder, DEP = Depression, SCZ = Schizophrenia, DBP = Diastolic blood pressure, T2D = Type 2 diabetes, RNA = ribonucleic acid, XWAS = both transcriptome and proteome wide association studies, MR = Mendelian randomization, coloc = colocalization.

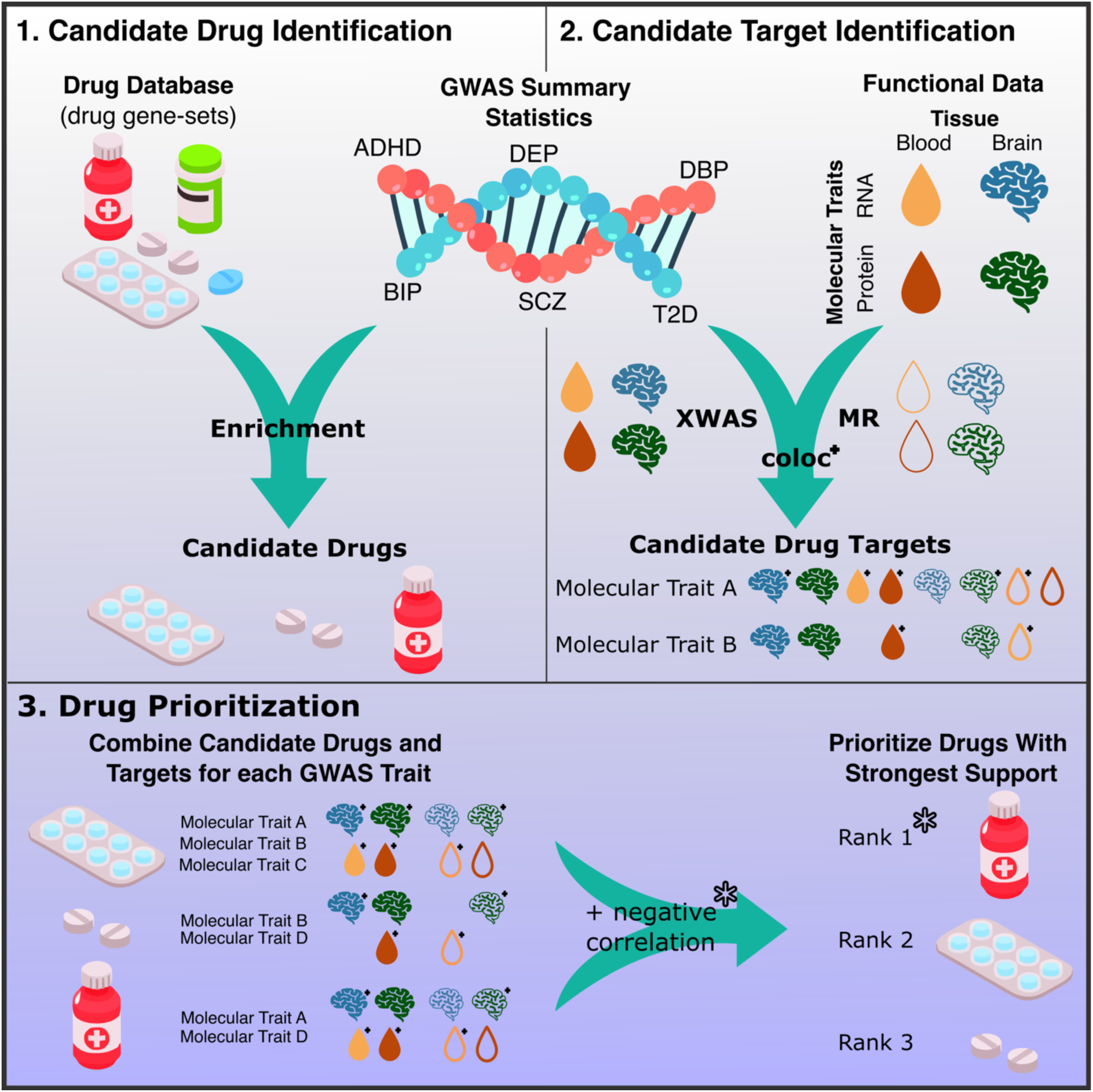

## Introduction

Genetics can provide insights into the underlying mechanisms of a trait that can be leveraged to inform drug development. Drugs with supporting genetic evidence have a 2.6 times greater probability of success in development compared to drugs lacking genetic support [1]. Notably, drugs for psychiatric treatment tend to have targets with less supporting genetic evidence than drugs for respiratory, metabolic, and cardiovascular diseases [1,2]. This lack of genetically informed drug development may partly explain the poor therapeutic response, adverse drug effects, and poor treatment compliance observed in psychiatry [1,3,4]. Furthermore, complex traits, such as psychiatric disorders and other medical conditions, share associated genes to a varying degree, a concept known as pleiotropy [5]. This pleiotropy may facilitate the repurposing of drugs approved or previously investigated for different indications. Therefore, leveraging genetics to inform drug discover, target prioritization, and repurposing may help improve psychiatric treatment.

The genome-wide association study (GWAS) of various psychiatric disorders have previously been leveraged to prioritize drug targets and compounds. Some studies use GWAS to perform gene-set analyses testing for enrichment of genes encoding drug targets [6–8]. While this approach has provided genetic support for various drugs with psychiatric indications, standard tools may be biased towards drugs with many targets potentially providing less disorder specificity [9]. Moreover, disorder-associated genetic variants can modulate gene expression and protein abundance providing a link to molecular traits (i.e., genes and proteins) serving as potential drug targets [10]. Therefore, transcriptome or proteome wide association studies (i.e., TWAS or PWAS) have been used to prioritize molecular traits as potential drug targets for several psychiatric disorders [11,12]. Moreover, Mendelian randomization (MR) is commonly used to identify causal relationships between molecular traits and psychiatric disorders [13–16]. Applying these analyses has revealed for example that calcium signalling is associated with multiple psychiatric disorders [7,11,14]. However, many studies tend to focus on one type of molecular trait or neglect bidirectional MR analyses. Also, while transcriptomic data provides greater coverage of the genome, drug targets are often proteins. Therefore, leveraging both types of molecular data is complementary and more comprehensive. Although studies leveraging multiple methodologies and types of molecular data to inform novel drug targets and indications exist [17–21], few integrate enrichment and comprehensive molecular trait analyses with a focus on psychiatric disorders.

Here, we present an extensive genetic interrogation of potential drug targets and compounds for attention deficit hyperactivity disorder (ADHD), bipolar disorder (BIP), depression (DEP), schizophrenia (SCZ) and as comparators two traits among clinical domains with strong biological support for medication development: diastolic blood pressure (DBP) and type 2 diabetes (T2D). We conduct gene-set analysis incorporating the novel GSA-MiXeR tool which identifies enrichment for biologically specific and smaller gene-sets than common approaches [9]. We use large scale molecular trait data from brain and blood tissue to identify potential drug targets. Blood is more readily available than brain tissue, resulting in larger datasets, and provides complementary identification of potential drug targets. We apply a series of molecular trait analyses including the novel use of UK Biobank (UKB) data for PWAS of psychiatric disorders. Finally, we combine results to prioritize potential drug targets and compounds.

## Materials and Methods

### Genome Wide Association Study Summary Statistics

We used summary statistics from the GWAS of four major psychiatric disorders in European samples. For ADHD we used a GWAS containing a total of 38,691 cases and 186,843 controls [22]. For BIP we used a GWAS including 41,917 cases and 371,549 controls [7]. For DEP we used a GWAS by Als et al. (2022) that excluded the 23andMe sample (n_cases_=294,322 n_controls_=741,438) [23]. For SCZ we used a GWAS including 76,755 cases and 243,649 controls [24]. As comparators, we used GWAS summary statistics for DBP containing a total of 1,076,093 individuals [25] and T2D containing 80,154 cases and 853,816 controls [26]. To avoid sample overlap in analyses with the UKB proteomic samples, we used summary statistics which excluded the UKB sample for BIP (n_cases_=40,463, n_controls_=313,436) and DEP (n_cases_=166,773, n_controls_=507,679). GWASs conducted by the Million Veterans Program were used for DBP (n=220,387)[27] and T2D (n=1,042,540)[28] to avoid sample overlap with blood molecular trait datasets. We chose DBP and T2D as comparators for several reasons: (i) DBP and T2D are not brain-related disorders and provide a non-psychiatric comparison for our approach to drug target and compound prioritization, (ii) in contrast to psychiatric medications, those developed for cardiovascular and metabolic conditions have more genetic support [1,2], and (iii) DBP is a continuous trait with summary statistics (e.g., betas) differing from the dichotomous disorder GWAS (e.g., odds-ratio/log odds-ratio) and (iv) both DBP and T2D have well powered publicly available GWAS summary statistics. These differences serve to enrich the assessment of our approach.

### Candidate Drug Identification

To obtain drug-target genes, we used data from the Drug Gene Interaction database (DGIdb; https://www.dgidb.org/)[29] which collates data from 28 different sources and is a common resource for drug enrichment analyses [6,7,30,31]. The Supplementary Material contains data processing details.

As our primary method to test each GWAS trait (i.e., ADHD, BIP, DEP, SCZ, DBP, T2D) for drug gene-set enrichment, we used GSA-MiXeR (https://github.com/precimed/gsa-mixer) [9]. GSA-MiXeR models gene-level heritability enabling the estimation of heritability partitioned across gene sets and fold enrichment which estimates the extent to which heritability is concentrated in a given gene set (Supplementary Material). An accompanying delta AIC value is used to indicate model fit where a positive value suggests good model fit. Trait-drug pairs with a positive delta AIC value were considered enriched. We additionally applied a gene-set analysis with MAGMA [32]. While MAGMA tends to identify enrichment among larger gene-sets than GSA-MiXeR [9], our approach requires observed enrichment with GSA-MiXeR and nominal significance (p<0.05) with MAGMA. This approach provides robust and potentially clinically actionable associations. All drug gene-sets with two or more genes were included in analyses.

A list of candidate drugs for each GWAS trait was determined based on enrichment using both GSA-MiXeR (AIC_delta_ >0) and MAGMA (p<0.05) and ranked using fold enrichment values from GSA-MiXeR. To assess the degree of overlapping enriched drugs between GWAS traits, we used a hypergeometric test. Enriched drugs were assigned an anatomic therapeutic chemical (ATC) code using classifications released in June 2023 from BioPortal. To determine if Level 1 ATC codes were over-represented among the candidate drugs for each GWAS trait, we used hypergeometric tests.

### Candidate Target Identification

#### Transcriptome- and Proteome-Wide Association Studies

To identify potential drug targets for each GWAS trait, we used the FUSION tool to conduct a TWAS and PWAS, which we jointly refer to as XWAS [33]. For TWAS using brain tissue, we used pre-computed single nucleotide polymorphism (SNP) weights for 14,751 genes from the PsychENCODE sample of 1321 individuals [34]. For TWAS using blood tissue, we used pre-computed weights for 8,125 genes from the genotype tissue expression (GTEx; n=558) resource [33,35]. For PWAS using brain tissue, we used pre-computed weights for 2,745 proteins in 720 participants from the Wingo et al. (2023) study [36]. For PWAS using blood tissue, we derived our own weights for 2841 proteins in 33,239 participants from the UKB (Supplementary Material). Since the T2D GWAS includes UKB participants, we used precomputed weights for the atherosclerosis risk in communities (ARIC: n=7,213, n_proteins_=1,348) sample and a separate GWAS which excluded the ARIC sample [37]. We performed correction for multiple comparisons across all XWAS analyses using the Benjamini-Hochberg method (q<0.05).

#### Mendelian Randomization

To further interrogate potential drug targets, we applied MR analyses using the R package TwoSampleMR [38]. For each trait, we performed analyses using local (cis) expression or protein quantitative trait loci (i.e., eQTL or pQTL), generally referred to as xQTLs. We assessed both forward causal relationships (i.e., from molecular trait to GWAS trait) and reverse causal relationships (i.e., from GWAS trait to molecular trait). The Supplementary Material provides details on the handling of potentially pleiotropic and proxy SNPs. Commonly, a single cis-xQTL remained after clumping therefore, the Wald ratio MR method was used. When multiple cis-xQTLs remained after clumping the inverse variance weighted approach was used.

From brain tissue, we used eQTL data for 18,397 genes from the MetaBrain sample (n=6,518)[39] and pQTL data for 7,553 proteins from the Religious Orders Study (ROS)/Rush Memory and Aging Project (MAP) sample (n=376)[40]. From blood tissue, we used eQTL data for 19,250 genes from eQTLGen sample (n=31,684)[41] and pQTL data for 2938 proteins from the UKB sample (n=34,090)[42]. To avoid sample overlap between eQTLGen with DBP and T2D, we used GTEx v8 eQTL data. In addition, to avoid sample overlap between T2D and UKB, we used the ARIC sample. We performed correction for multiple comparisons across all MR analyses using the Benjamini-Hochberg method (q<0.05).

#### Colocalization

Colocalization analyses help determine if two phenotypes share causal SNPs. For all nominally significant (p<0.05) molecular trait associations, we conducted colocalization analyses with the associated GWAS trait using the R package COLOC [43]. We considered the molecular and GWAS trait colocalized when the posterior probability of colocalization was greater than 0.8 (Supplementary Material).

#### Prioritization of Candidate Targets

To generate a ranked list of candidate drug targets for each GWAS trait, we combined evidence across the molecular trait analyses (i.e., XWAS, MR, and colocalization). For each molecular trait analysis (e.g., brain TWAS or blood pQTL colocalization) we normalized association statistics (e.g., Z-scores or PP.H4). Normalized values were generated by dividing the absolute value of the association statistic by the sum of the absolute values across all molecular traits. The resulting normalized values fall between 0 and 1 with larger values representing stronger associations between molecular and GWAS traits. Next, for each molecular trait, the mean normalized value across all analyses, excluding missing values, was used as an overall score. Therefore, molecular traits with the strongest average GWAS associations rank highly without penalization for a lack of representation. A brain-score was generated using the mean normalized values from all brain-tissue analyses. Then, a brain-weighted score was generated as the product of the overall and brain scores. The brain-weighted score provides tissue specificity while also leveraging the large statistical power of blood tissue analyses. This final brain-weighted score was used to rank molecular traits for a given GWAS-trait. For each GWAS trait, we performed a gene-ontology analysis of these candidate targets using the R package clusterProfiler.

### Drug Prioritization

#### Negative Correlation Test

We extracted drug-induced gene expression data from Connectivity Map 2020 for all enriched drugs using the phase 2 release of the Library of Integrated Cellular Signatures (LINCS) with the cmapR package (v 4.3.1) [44,45]. For each drug and GWAS trait pairing, a Spearman correlation between drug induced gene expression and variation in molecular trait associations was conducted. Negative correlations indicate the drug may reverse changes in molecular trait abundance associated with the GWAS trait. This “negative correlation test” was run separately for each XWAS and MR analyses and corrections for multiple comparisons were conducted using the Benjamini Hochberg method (q<0.05).

#### Prioritization of Drugs

To prioritize drugs for each GWAS trait, we calculated the mean brain-weighted score across drug-target genes. Therefore, drugs with the strongest molecular trait support were ranked highest. Then a final drug score was generated as the product of the mean brain-weighted score and the normalized fold-enrichment values from GSA-MiXeR. This final score was used to rank drugs for a given GWAS trait. If a negative correlation test was observed for a given drug and GWAS-trait pairing, that drug was moved one place up the ranking given the added supporting evidence.

#### Comparison of Drugs with Variable Efficacy

We assessed genetic support for a list of common antidepressants, antipsychotics, mood stabilizers/anticonvulsants, and stimulants (Supplementary Material). In addition, we assess genetic support for a selection of drugs with failed clinical trials for medications designed to treat psychiatric disorders (Supplementary Material). Here, we assessed the molecular trait support for each drug irrespective of whether the drug was enriched using GSA-MiXeR.

### Ethics

This study was approved by the Regional Committee for Medical Research Ethics including the use of individual and genetics data from the UKB (accession number 27412).

## Results

### Drug Enrichment Analyses

We used 9,460 drug gene-sets (Supplementary Table 1) to perform enrichment analyses. GWAS of ADHD, BIP, DEP, and SCZ were enriched for a total of 12, 107, 113, and 202 drugs using both GSA-MiXeR (n=34, 152, 283, and 295, respectively; Supplementary Table 2) and MAGMA (n=400, 686, 566, and 713, respectively; Supplementary Tables 3). We observed substantial overlap among the candidate drugs for BIP and SCZ (n=63, p= 3.51e-11) but less among the other psychiatric disorders (BIP-DEP: n=5, p=1.00, DEP-SCZ: n=13, p=1.00) and ADHD only had one overlapping drug with DEP (Figure 1). The ATC code for nervous system drugs (Supplementary Table 4) was enriched, for BIP (n=12/35, p=1.49e-8) and SCZ (n=41/92, p=1.27e-21) but not ADHD (n=1/6, p=0.07) or DEP (n=7/72, p=9.65e-1). Among the top 10 candidate drugs for BIP, DEP, and SCZ were several drugs with antipsychotic properties (e.g., adoprazine, benperidol, cariprazine, mazapertine, nemonapride, piperacetazine, sarizotan, and stepholidine). For ADHD, the top 10 candidate drugs were all either pro- or anti-cholinergic drugs.

**Figure 1.**
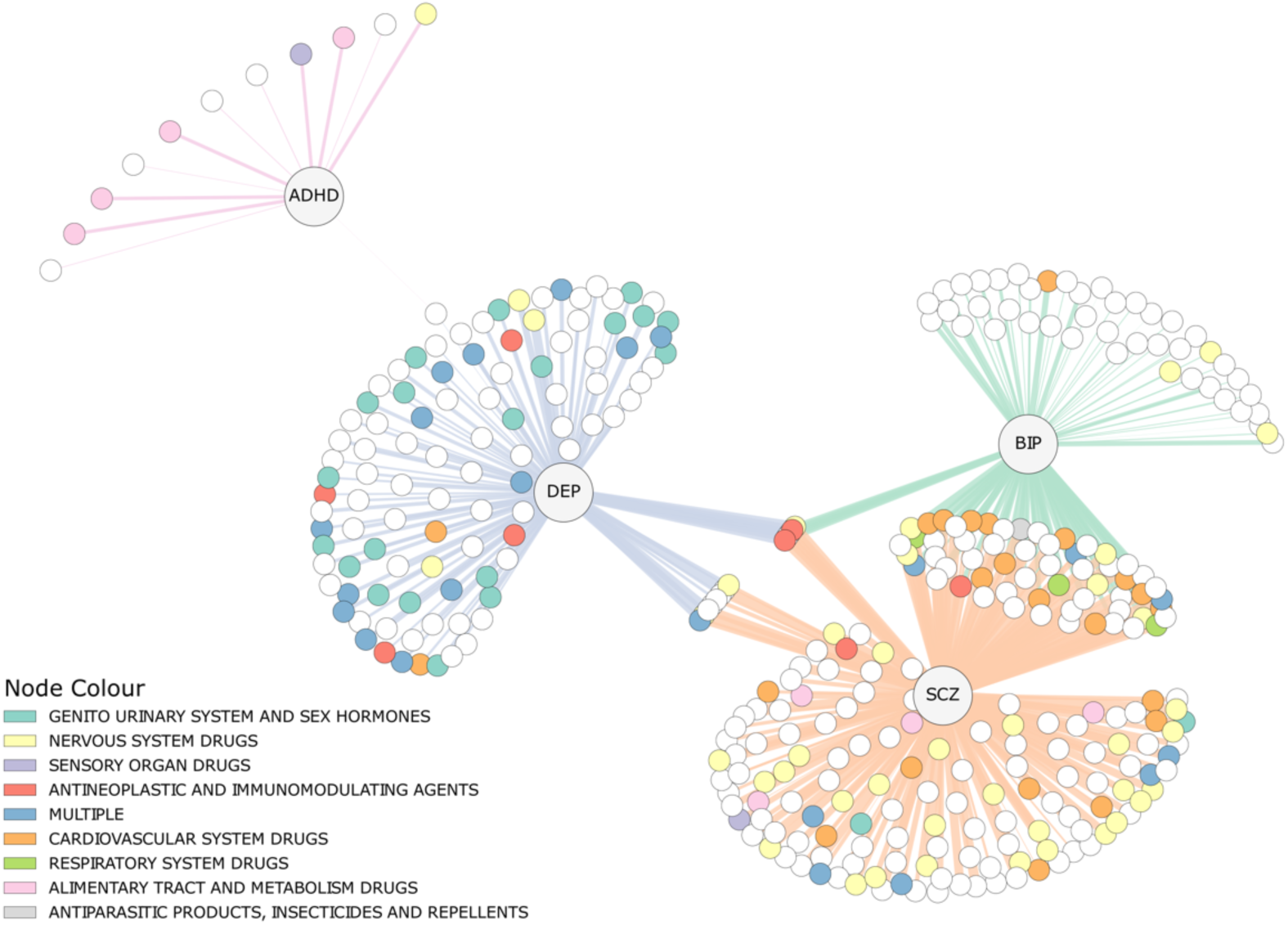
Overlap of Enriched Drugs for Psychiatric Disorders. A network plot depicting each of the enriched drugs (colored nodes) for each psychiatric disorder labelled central hubs. Each drug node is colored based on their level 1 anatomical therapeutic chemical classification. Those nodes without a color (white) represent drugs not assigned an anatomical therapeutic chemical code. The thicker the edge (line connecting nodes to disorder), the larger the fold enrichment. ADHD = Attention deficit hyperactivity disorder, BIP = Bipolar disorder, DEP = Depression, SCZ = Schizophrenia, MULTIPLE = A drug with multiple classifications.

As for the comparators, DBP was enriched for 257 drugs and T2D for 95 (n_GSA-MiXeR_=517 and 273, respectively; n_MAGMA_=1029 and 479, respectively; Supplementary Tables 2 and 3). No significant overlap between candidate drugs for DBP or T2D and the psychiatric disorders was observed (Supplementary Material Figure 1). There was no ATC code enrichment for nervous system drugs (DPB: n=2/163, p=1.00; T2D: n=2/43, p= 9.47e-1) but DBP had enrichment for cardiovascular system drugs (n=80/163, p=5.86e-35) and T2D for alimentary tract and metabolism drugs (n=21/43, p=2.42e-22).

### Molecular Trait Analyses

XWAS analyses identified 504, 1142, 1160, and 1855 significant molecular trait associations for ADHD, BIP, DEP, SCZ, respectively (Figure 2, Supplementary Table 5). The molecular traits associated with enriched drugs are labelled in Figure 2 where several were supported by multiple XWAS analyses such as *ANKK1* with DEP. Colocalization analyses revealed 935 (20.06%) of the significant XWAS associations with psychiatric disorders had colocalized genetic signal (Supplementary Tables 6) with enriched drug-target genes depicted in Figure 2 with an asterisk. No significant negative correlation test was observed after correction for multiple comparisons (Supplementary Table 7).

**Figure 2.**
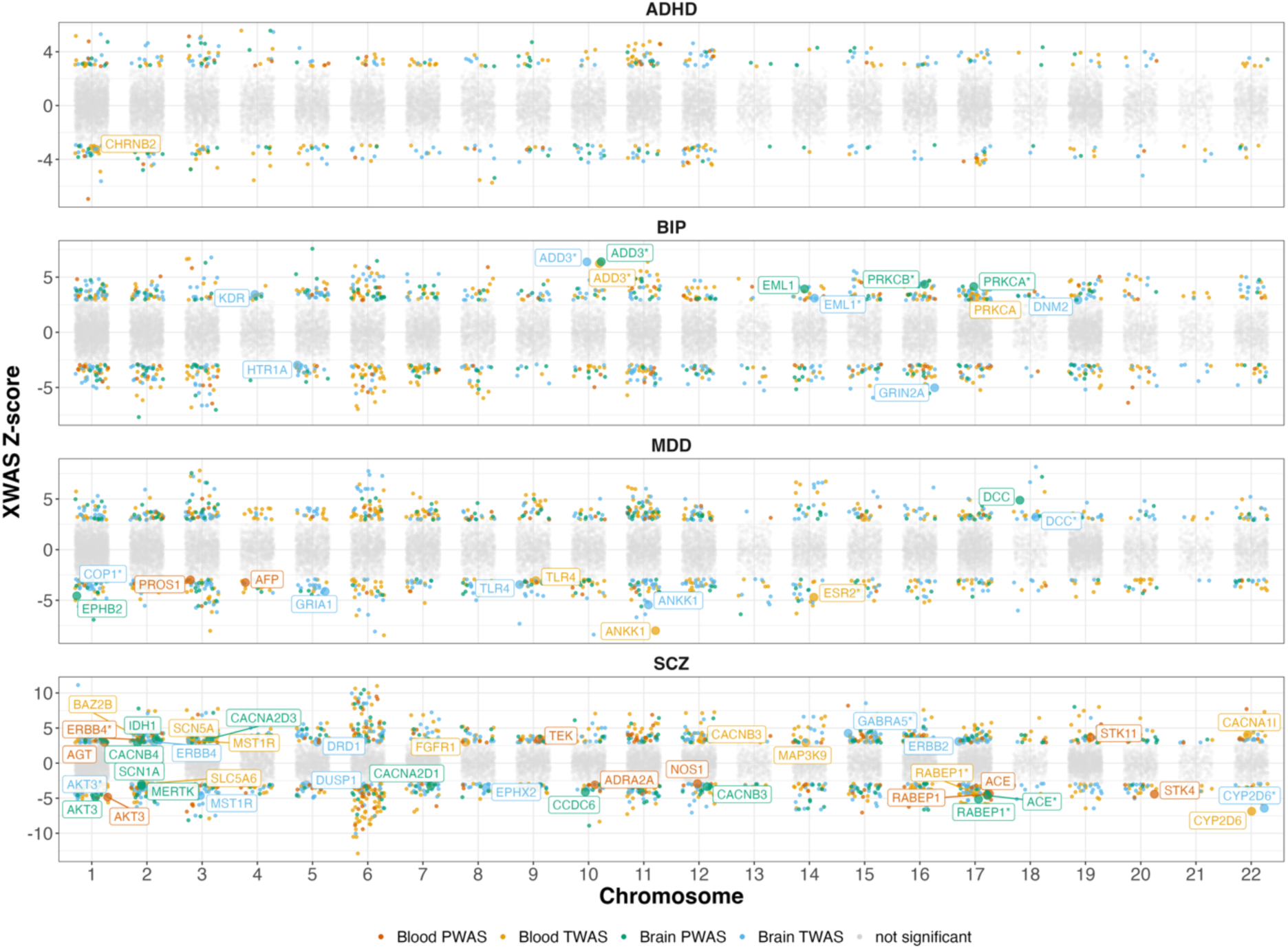
Transcriptome- and Proteome-Wide Association Study Results. For each psychiatric disorder the results of transcriptome- and proteome-wide association studies are presented. Significantly associated molecular traits that are also a part of gene sets for enriched drugs are labelled. Labelled molecular traits that were also colocalized are represented with an asterisk. ADHD: attention deficit hyperactivity disorder, BIP: bipolar disorder, DEP: Depression, SCZ: schizophrenia, TWAS = Transcriptome wide association study, PWAS = Proteome wide association study.

Across the bidirectional MR analyses, 588, 1344, 1462, and 2421 molecular traits were significantly associated with ADHD, BIP, DEP, and SCZ, respectively (Figure 3, Supplementary Table 8). The molecular traits associated with enriched drugs are labelled in Figure 3 where several were supported by multiple MR analyses such as *ADD3* with BIP and *AKT3* with SCZ. Colocalization analyses revealed 694 (11.93%) of the significant MR associations had colocalized genetic signal (Supplementary Tables 9) and with enriched drug-target genes depicted in Figure 3. There were no significant negative correlation tests (Supplementary Tables 10).

**Figure 3.**
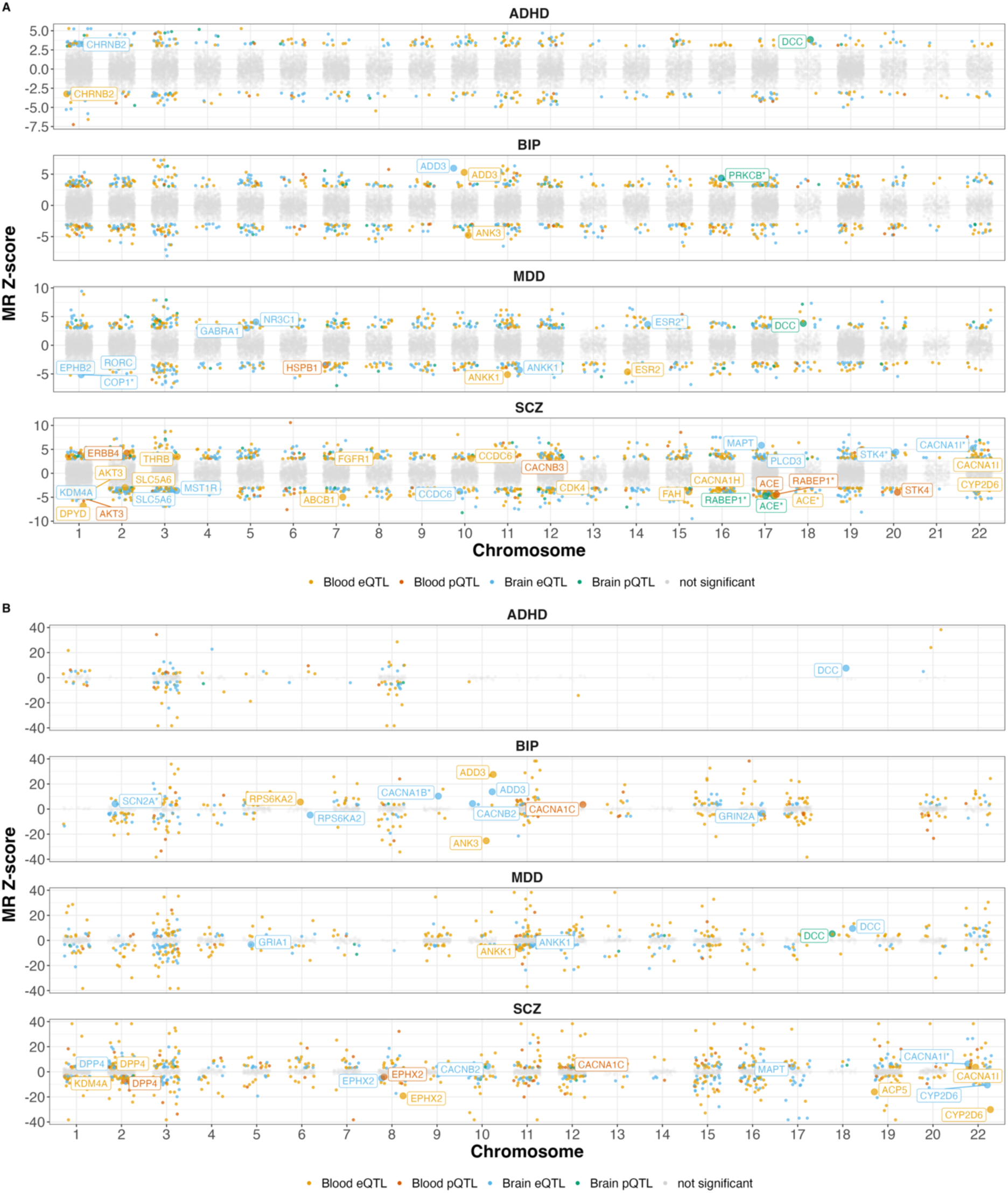
Mendelian Randomization Results. For each psychiatric disorder the results of the Mendelian Randomization (MR) analyses are presented. Panel A shows the MR results in the forward direction with the psychiatric disorders as the outcome traits. Panel B shows the MR results in the reverse direction with the psychiatric disorders as the exposure traits. Significantly associated molecular traits that are also a part of a drug gene set enriched for the disorder are labelled. Labelled molecular traits that were also colocalized are represented with an asterisk. ADHD: attention deficit hyperactivity disorder, BIP: bipolar disorder, DEP: Depression, SCZ: schizophrenia, eQTL = expression quantitative trait loci, pQTL = protein quantitative trait loci.

DBP had 3707 significant XWAS associations and 3112 MR while T2D 1500 significant XWAS and 1799 MR associations (Supplementary Tables 5 and 8) with many enriched drug-target genes implicated (Supplementary Material Figures 2 and 3). Colocalization was revealed for 614 (16.56%) and 330 (22.00%) XWAS as well as 253 (8.13%) and 186 (10.34%) MR associations for DBP and T2D, respectively (Supplementary Tables 6 and 9). No significant negative correlation test was observed (Supplementary Tables 7 and 10).

### Prioritization of Potential Drug Targets and Compounds

We identified 795, 1657, 1720, and 2650 candidate drug targets supported by at least one molecular trait analysis for ADHD, BIP, DEP, and SCZ, respectively (Supplementary Table 11-14). Among these candidate targets, there was significant overlap observed for BIP with SCZ (n=661, p= 1.79e-19) and DEP (n=359, p=0.02) as well as ADHD and DEP (n=219, p= 2.39e-8). Figure 4 presents the top 10 candidate targets for each psychiatric disorder and their supporting evidence. The gene ontology term “glutamatergic synapse” was enriched among the candidate targets for BIP, DEP, and SCZ (Supplementary Table 17). DBP and T2D had many candidate targets (n_DBP_=4310, n_T2D_=2191; Supplementary Table 15-16) and some shared gene ontology terms with psychiatric disorders such as “glutamatergic synapse”, “MHC protein complex”, and “early endosome”.

**Figure 4.**
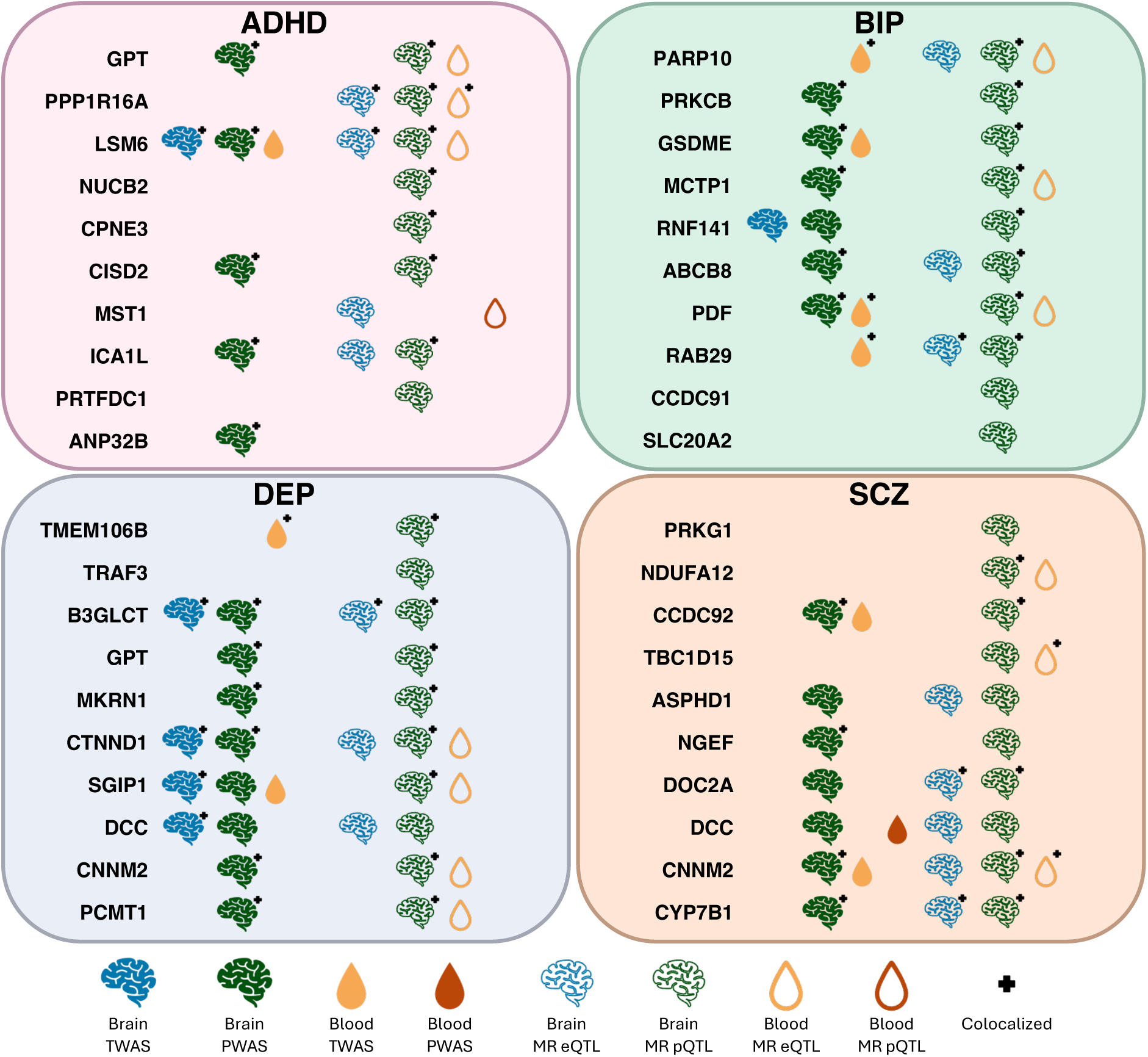
Prioritization of Candidate Drug Targets. The list of prioritized potential drug targets for each psychiatric disorder after combining results of molecular trait analyses from transcriptome and proteome wide association studies, Mendelian randomization, and colocalization. ADHD: attention deficit hyperactivity disorder, BIP: bipolar disorder, DEP: Depression, SCZ: schizophrenia, TWAS = Transcriptome wide association study, PWAS = Proteome wide association study, MR = Mendelian randomization, eQTL = expression quantitative trait loci, pQTL = protein quantitative trait loci.

After combining molecular trait support with drug enrichment for each psychiatric disorder, 2 (16.67), 67 (62.62 %), 65 (57.52%), and 116 (57.43%) drugs were supported for ADHD, BIP, DEP, and SCZ, respectively (Supplementary Table 18-21). Figure 5 presents the top 10 prioritized candidate drugs and their supporting evidence. For ADHD, a matrix metalloproteinase inhibitor and acetylcholine receptor agonist were enriched drugs with molecular trait support. For BIP, a mixture of calcium channel blockers, protein kinase inhibitors, and drugs with antipsychotic/antiepileptic properties were top ranked. For DEP, the top 10 drugs included predominantly estrogen related compounds except the top two ranked matrix metalloproteinase inhibitors. For SCZ, seven antipsychotics were among the top 10 ranked drugs.

**Figure 5.**
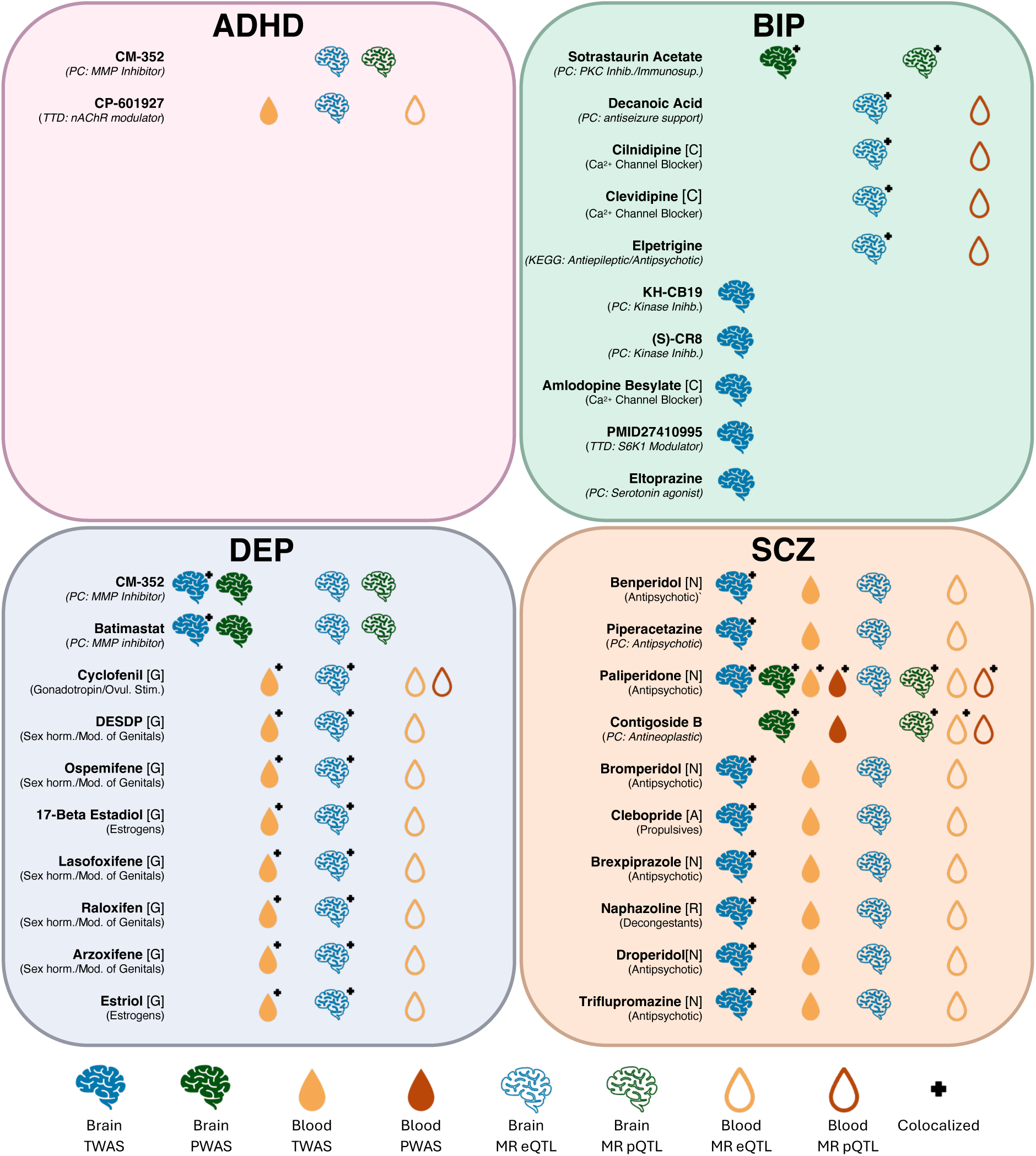
Prioritization of Drug Compounds. The prioritized list of drug compounds (in bold) for each psychiatric disorder with molecular trait support for enriched drugs. In brackets next to each drug compound is the level 1 anatomical therapeutic chemical (ATC) classification code. Below each drug compound is a brief description. In most cases the description is based on the level 3 ATC classification. Drugs without an ATC code use descriptions from various other sources including: PubChem (PC; https://pubchem.ncbi.nlm.nih.gov/), Kyoto encyclopedia of genes and genomes (KEGG; https://www.genome.jp/kegg/kegg2.html), Therapeutic target database (TTD; https://idrblab.net/ttd/). The drug KH-CB19 is a synonym for ethyl 3-[(e)-2-amino-1-cyanoethenyl]-6,7-dichloro-1-methyl-1h-indole-2-carboxylate which is the name used in supplementary materials. nAChR = nicotinic acetylcholine receptor, MMP = Matrix metalloproteinase, DESDP = Diethylstilbestrol Diphosphate, [C] = Cardiovascular system drug, [G] = Genito urinary system and sex hormones, [N] = Nervous system drug, PKC inhib/immunosup. = Protein kinase C inhibitor and immunosuppressant.

DBP and T2D enriched drugs had strong molecular trait support [n_DBP_=220 (85.60%) and n_T2D_=64 (67.37%)] (Supplementary Table 22-23). The top 10 candidate drugs for DBP included several approved antihypertensives (e.g., ilepatril, deserpidine, and quinapril). For T2D, the top 10 candidate drugs included several approved for treatment (e.g., repaglinide, mitiglinide, tolbutamide, acetohexamide, and gliclazide).

Analyses assessing molecular trait support for common psychotropics (Supplementary Table 24) revealed: (i) common stimulants used to treat ADHD, (e.g., methylphenidate) had no molecular trait support, (ii) DEP had molecular trait support for several antidepressants (e.g., duloxetine, clomipramine, and citalopram) but was never the top ranked GWAS trait, (iii) SCZ had molecular trait support and was the top ranked GWAS trait for 10 (e.g., olanzapine, paliperidone, risperidone, and clozapine) antipsychotics followed by BIP then DEP, (iv) BIP had support for several mood stabilizers/anticonvulsants and was the top ranked GWAS trait for lamotrigine (Supplementary Table 24). Among the psychotropics with failed clinical trials (Supplementary Table 25), there was either no molecular trait support for the outcome disorder of the trial (e.g., metadoxine for ADHD, esmethadone for DEP or ulotaront for SCZ), or a low level of support (e.g., brexpiprazole for BIP).

## Discussion

We prioritized potential drug targets and compounds for psychiatric disorders using an extensive genetically informed investigation. Our approach employed novel tools such as GSA-MiXeR [9] for drug gene-set enrichment and the novel use of UKB proteomic data [42] for PWAS of psychiatric disorders. The series of molecular trait analyses revealed candidate drug targets with varying degrees of overlap between the psychiatric disorders. The prioritization of potential drugs revealed known and investigational compounds with indications for psychiatric treatment as well as potential opportunities for repurposing. Altogether, this work highlights the potential utility of genetics for informing psychiatric drug development.

Top-ranked candidate targets exhibited varying degrees of overlap among the psychiatric disorder and were consistent with previous reports. DCC was among the top ranked candidate targets for DEP and SCZ with previous genetic links to psychiatric disorders [24,46,47]. DCC plays a crucial role in axonal growth during brain development including dopaminergic pathways and developmental disruptions may result in brain structural dysconnectivity linked to various psychiatric disorders [48–50]. Both ADHD and DEP had GPT, an enzyme involved in metabolism of glucose and amino acids such as glutamate, among their top ranked candidate targets which provides a potential link to energy metabolism and glutamate associated neurotransmission. Moreover, sharing of candidate targets between BIP, DEP, and SCZ revealed enrichment for the gene ontology term “excitatory glutamatergic synapse” providing further support for glutamate signalling as a therapeutic target for psychiatric disorders and suggests the need for more therapeutic investigations of glutamate signalling in psychiatry [51,52]. Meanwhile, we and others [12,53] have linked B3GLCT to DEP and since we found only BIP was also associated, B3GLCT may be mood disorder specific. While these findings require further investigation of a wider variety of disorders, revealing potential drug targets specific to a subclass of disorders can be useful for targeting similar cross-disorder symptomology or commonly co-morbid disorders. Many of our top ranked candidate targets have been previously linked to their associated psychiatric disorders in the original GWAS or through various molecular trait analyses. Examples of these associations include: PPP1R16A and LSM6A with ADHD [22]; CNNM2, CTNND1, and MKRN1 with DEP [12,23,54]; PDF, PRKCB, and ABCB8 with BIP [54]; ASPHD1, DCC, CNNM2, and CYP7B1 with SCZ [24,54]. The consistency across reports further strengthens the support for the prioritized candidate targets.

Consistent with previous studies, our results provide genetic support for drugs with known or putative indications for psychiatric treatment [6,7,11,31]. In particular, the top 10 prioritized drugs for SCZ included 7 antipsychotics. In addition, we find that drugs with failed clinical trials either have no molecular trait support for their indicated disorder or were ranked low on the list of prioritized drugs. These findings help validate our approach for prioritizing drugs. However, our findings also reveal a lack of genetic support for stimulants for treating ADHD [55]. Notably, ADHD had the lowest drug enrichment and molecular trait associations which likely reflect the lower power of the GWAS compared to the other psychiatric traits. Therefore, with future larger GWAS, a clearer picture of the genetic architecture of psychiatric disorders will be revealed and can be further leveraged to inform drug development.

Identified drugs in our study additionally highlight opportunities for repurposing. The majority of enriched drugs for ADHD modulate acetylcholine receptors and one agonist (CP-601927) had candidate target support, which is in line with the previously proposed use of cholinergic drugs for ADHD treatment [56]. Top ranked drugs for ADHD and DEP included matrix metalloproteinase (MMP) inhibitors which have been proposed as a new avenue for drug development and repurposing in psychiatrics [57]. MMPs are involved in synaptic remodelling and inflammatory responses in the brain and have been linked to various psychiatric disorders [57]. After MMP inhibitors, the top ranked drugs for DEP were predominantly estrogen modulators which have previously been linked to DEP but are not currently indicated treatment [58–60]. While estrogen is a female sex hormone it’s modulation as a putative therapeutic target may be relevant for both males and females [61–63]. More work is needed to understand the sex-specific genetic support for psychiatric drug development where sex-stratified analyses are critical.

Combining enrichment with molecular trait analyses revealed important clinical implications for enriched drug targets. Across several analyses and consistent with previous reports, genetic liability to SCZ was associated with reduced expression of *CYP2D6*, an enriched drug target that plays a role in metabolism of neurotransmitters and drugs, including antipsychotics [14,64–66]. Therefore, lower levels of *CYP2D6* may contribute to SCZ pathophysiology and also disrupt treatment response which supports efforts for therapeutic monitoring of *CYP2D6* polymorphisms and evidence-based drug dosing [14,67]. ANKK1 was negatively associated with DEP in several analyses, includes a polymorphism linked to reductions in dopamine receptor abundance, and is a target for several psychotropic drugs [68,69]. Therefore, while many antipsychotics have high affinity for both dopamine and serotonin receptors [70], ANKK1 may be an alternative link for response to antipsychotics in patients with DEP. Consistent with previous reports, BIP and SCZ were associated with candidate targets related to cellular calcium supporting the prioritization of calcium channel blockers for BIP and their continued investigation as therapeutic drugs in psychiatry [7,14,71]. Altogether, these current drug targets revealed important implications for drug development.

This study has several strengths and limitations. Using DBP and T2D as comparators revealed a lack of overlap in candidate drug targets and enriched drug compounds with any psychiatric disorder and showed the generalizability of our approach beyond psychiatric disorders. A limitation to our MR analyses is the potential for weak instrument bias particularly when using a single instrument approach. This may bias results towards the null leading to a failure to detect true relationships between molecular traits and GWAS traits [38]. Our approach prioritizes drugs irrespective of effect direction. Therefore, we identify drugs with genetic evidence for beneficial or detrimental effects for a trait. While this is useful, our findings require careful interpretation and further investigation. We also use transcriptomic and proteomic data from both brain and blood tissue. The molecular traits and tissue sources are complimentary but not completely correlated.

Nevertheless, their integration improves the power to identify modifiable drug targets [72,73]. Our findings may lack generalizability beyond individuals of European ancestry due in part to the reliance of tools on European references and a lack of well powered non-European GWASs and sources of molecular trait data. Additionally, the power of the included GWAS traits vary, which likely contributes to greater drug enrichment and molecular support for GWAS traits with higher power (i.e., SCZ and DBP) compared to those with lower power (i.e., ADHD). This power discrepancy also effects the observed overlap across GWAS traits.

In conclusion, our extensive analytical approach leveraged psychiatric GWAS combined with novel tools and molecular trait data to prioritize potential drug targets and compounds. The candidate targets can be used for future development of psychotropics. Meanwhile, the prioritized drugs can be used for future repurposing efforts. This study provides clinically relevant evidence for how psychiatric genetics can inform future drug development and repurposing with the potential to improve patient outcomes.

## Data Availability Statement

All GWAS summary statistics and molecular trait data included in this study are publicly available. We generated PWAS weights using proteomic data from the UK Biobank and have shared those weights on figshare [https://doi.org/10.6084/m9.figshare.31487881]. The GWAS summary statistics from the Million Veteran Program were downloaded from the dbGaP web site, under accession phs001672.

## Acknowledgments

We thank all participants, staff, and providers of the publicly available data used in this study including individuals from the UKB, psychiatric genomics consortium, Million Veterans Program, PsychENCODE, ARIC, MetaBrain, ROS/MAP, eQTLGen, GTEx.

## Author Contributions

NP and OBS conceived of the study and EK, AAS, GFLH, SS, PJ, KSO, and OAA were involved in study design. NP, EK, AAS, TSW, and APW were involved in data acquisition and processing. NP, EK, AAS, and JF were involved in data analysis. NP had access to all data and drafted the initial manuscript. All authors contributed to data interpretation and editing of the manuscript and accepted responsibility to submit the manuscript for publication.

