## Supplementary Methods and Figures for "Leveraging the Genetics of Psychiatric Disorders to Prioritize Potential Drug Targets and Compounds"

#### Generating Drug Gene Sets

The Drug Gene Interaction database (DGIdb) was used to acquire genes associated with various drug compounds. We used the DGIdb provided “drug name”, if missing we used “drug claim name”, for each drug compound. We removed duplicated genes associated with a given compound. It is this list of genes associated with each drug that was used to construct the drug support scores when compiling evidence across molecular trait analyses. However, for the enrichment analyses with GSA-MiXeR [1] and MAGMA [2], gene sets were restricted to the set of 18,201 protein coding genes defined by the GSA-MiXeR analyses. Additionally, only drug gene-sets that had two or more genes were included to comply with the MAGMA analyses.

#### Gene Set Analysis MiXeR (GSA-MiXeR) Fold Enrichment

The GSA-MiXeR method is designed to estimate enrichment for a particular gene set for a given trait of interest using genome-wide association study (GWAS) summary statistics. To do so, the tool models gene-level heritability with associated error bars and estimates partitioned heritability and fold enrichment. To estimate fold enrichment two models are fit to the GWAS summary statistics. The full model estimates individual gene contributions to the heritability of a trait while the base model estimates heritability without accounting for the distribution of individual genes. The fold enrichment for a gene set is then considered as the ratio of heritability from the full model over the baseline model. In addition, the Akaike information criterion (AIC) for both the full and the base model are calculated. The difference in the AIC values, or the delta AIC, informs model fit and a positive delta AIC represents a well-fitting full model. Therefore, gene-sets with a positive delta AIC are considered enriched when using GSA-MiXeR.

#### Derivation of UK Biobank Protein Weights

##### *Genetic and Sample Data Quality Control*

We used genotype data provided by the UK Biobank through application number 27412. Using PLINK v1.9 [3], we performed both single nucleotide polymorphism (SNP) and individual/subject level quality control (QC) on the genetic data. The SNP QC included a minor allele frequency threshold of 0.01, Hardy Weinberg equilibrium threshold at 1e-10, missingness threshold of 0.05. For individual QC, we set a SNP missingness threshold of 0.01 and performed QC of heterozygosity rates by removing individuals with an F coefficient greater than 5 standard deviations from the mean. In addition, with non-imputed genetic data, we generated the first 20 genetic principle components and a list of unrelated individuals with a relatedness cut-off of 0.1. We additionally restricted the sample to (1) exclude participants who had dropped out of the UK Biobank study, (2) only include the baseline proteomics sample (excluding the consortium and COVID sample)[4], (3) remove individuals with a sex chromosome aneuploidy, and (4) only include individuals of white British ancestry.

##### *Proteomic Data*

We used normalized protein data previously generate by the UK Biobank [4]. The measures of protein abundance were pre-residualized for the following covariates using linear regression: age, age-squared, sex, age*sex, age-squared*sex, proteomic batch, genetic array, and the first 20 genetic principle components. Next, we generated two different sets of proteomic outputs to be used to generate protein weights for proteome-wide association study (PWAS) analyses, (1) residuals of the above model and (2) rank inverse normal transformed residuals from the above model. A third set of protein values were generated by first filtering out all provided normalized protein expression values that were below the limit of detection. Then the filtered data was pre-residualized using the same covariates listed above and rank inverse normal transformed. We performed all PWAS analyses using each of the three models. Notably, PWAS Z-values were highly correlated across the three analyses (r>0.85). Additionally, the proportion of associations which remained nominally significant across the 3 types of protein pre-processing was high (>97%). The main results presented in the text used the protein dataset #2 above (i.e., rank inverse normal transformed residuals without limit of detection filtering). We did this to include a larger number of proteins in the analysis, compared to limit of detection filtering. Also, most PWAS weight are derived using rank inverse normalized protein abundance values and the original proteomics UK Biobank paper did not conduct limit of detection filtering [4,5].

After processing the protein data, we used the FUSION tool [5] to generate weights for PWAS analyses. We generated weights using three models (i.e., top1, lasso, enet) provided by the FUSION tool. Similar to previous studies, we set our cis-region boundary to be within 1MB of the gene encoding the protein [6]. Those weights were then subsequently used for PWAS analyses. Of note, some of the assayed proteins can have sub-units comprised of multiple genes, either on the same chromosome or not. Therefore, for multi-gene proteins, we generated weights for individual sub-units. While the expression values for each subunit is the same, the SNPs in the cis-region (+/- 1MB from the gene body) vary based on the position of the coding gene. This can ultimately result in differing protein-to-trait associations in the proteome wide association study (PWAS) for each sub-unit.

To avoid attempts to calculate heritability for each protein, which is computationally intensive for large samples, we set heritability of each protein to 0.1. This allowed FUSION to proceed with analyses for all proteins without calculating heritability. Then, to ensure strong genetic signal was present within the cis-region (+/- 1mb) of included proteins, we restricted all results to proteins which had at least one genome-wide significant (p<5e-8) hit in the cis-region. This procedure is used to proxy heritable genetic signal for the protein in XWAS studies. The restriction resulted in a total of 2086 proteins included in the final PWAS results.

**Mendelian Randomization Data Processing and Analysis**

Mendelian randomization (MR) is a method to estimate causal associations between an exposure and an outcome. Genetic variants associated with the exposure are used as "instruments" for MR analyses. Here we perform MR analyses in two directions. In the first "forward" direction, gene expression or protein abundance are set as an exposure and psychiatric disorders or our comparator GWAS traits (i.e., DBP and T2D) are set as an outcome. This way we use cis-region expression and protein quantitative trait loci (eQTLs and pQTLs) as instruments. For the second "reverse" direction, psychiatric disorders (or comparator GWAS traits) are set as the exposure and gene expression and protein abundance are the outcome. Performing this bidirectional analyses helps to identify not only genes/proteins which may play a causal role in developing a psychiatric disorder (or comparator GWAS traits) but also the reverse causal associations where genes/proteins are altered as a result of the disorder. Identified genes/proteins in both directions can be potential drug targets.

We performed some additional processing of the UK Biobank pQTL data and the eQTLGen eQTL data. The UK Biobank perfromed a genome-wide analysis of proteomic data derived from several Olink panels [4]. Since all other datasets provided cis-QTLs, we restricted the summary statistics provided by the UK Biobank to the cis-region (+/- 1MB) of the coding gene. Some proteins were composed of more than one gene. In this case, the cis-region of each sub-unit was combined for the MR analysis. For the eQTLGen dataset, the summary statistics contained only Z-values as effect sizes. Meanwhile, MR analyses relies on using betas and standard errors. Therefore, we used a common method proposed by Zhu et al (2016) which estimates betas and standard errors using Z-values, minor allele frequencies, and sample size [7].

There are many methods to perform MR. However, our analyses with cis-eQTLs and cis-pQTLs restricts the number of independent variants that can be included in the MR analyses to as little as one instrument. Therefore, in cases where one instrument is available, we use the Wald ration approach and for cases where more than one instrument is available we use the inverse variance weighted approach all implemented in the TwoSampleMR package in R [8]. When performing analyses with genes/proteins as exposures, remove potentially pleiotropic instruments prior to analysis. Similar to previous studies [9,10], we define pleiotropic instruments as eQTLs/pQTLs from a given source dataset that are associated with five or more genes/proteins. We also use proxy variants with an $R^2$ of 0.8 or higher for instruments that are not present in the outcome summary statistics. We generated proxy variants using PLINK v1.9 [3].

**Details on Colocalization Analyses**

To provide additional evidence of trait associations with genes/proteins we applied a Bayesian colocalization analysis using the COLOC package in R [11]. We ran colocalization analyses for all XWAS and MR associations that were nominally significant ($p<0.05$).
We used the procedure outlined by the FUSION R package (http://gusevlab.org/projects/fusion/) to run COLOC for the XWAS analyses. The procedure estimates colocalization using the FUSION weights and assumes standard errors are equivalent to the inverse of the sample size. For associations identified by MR we ran COLOC using effect sizes and standard errors provided in the same summary statistics used for as the MR analyses (i.e., cis-eQTLs/cis-pQTLs and GWAS summary statistics).

We applied the approximate Bayes factor colocalization analyses to estimate the support for the following hypotheses:

$H_0$: neither trait has a genetic association in the cis-region
$H_1$: only the first trait has an association in the cis-region
$H_2$: only the second trait has an association in the cis-region

$H_3$: both traits are associated but with different causal variants
$H_4$: both traits are associated and share a causal variant

For each analysis probabilities are generated for each hypothesis. We use a probability greater than 0.8 for $H_4$ to represent colocalized genetic signal.

**Commonly used Medications**

**Antidepressants:** amitriptyline, bupropion, citalopram, clomipramine, duloxetine, fluoxetine, phenelzine, sertraline, tranylcypromine, venlafaxine
**Antipsychotics:** aripiprazole, chlorpromazine, clozapine, haloperidol, olanzapine, paliperidone, quetiapine, risperidone
**Mood stabilizers/Anticonvulsants:** lithium, carbamazepine, lamotrigine, valproic acid
**Stimulants:** dextroamphetamine, lisdexamfetamine, methylphenidate

**Psychotropic Drugs with Failed Clinical Trials**

Here we report a list of drugs with clinical trials which failed due to either a failure to meet primary endpoints, unmanageable toxicity/poor pharmacokinetics, and/or a lack of efficacy. The list includes drug for ADHD: metadoxine; for BIP: brexpiprazole and lumateperone; for DEP: NMRA-140, esmethadone; and for SCZ: xanomeline and ulotaront.

**Data Harmonization**

For all analyses performed in this study, we converted genetic data to genomic build hg37. We did so to harmonize datasets to the same genomic coordinates. The choice of hg37 was because most genetic summary statistics were provided in this build. The build conversions were done using the liftOver command line tool (https://genome.ucsc.edu/cgi-bin/hgLiftOver).

We also harmonized molecular trait data in order to combine supporting evidence for each GWAS trait. If gene data was provided using ENSMBL IDs we used the biomaRt package in R to map ENSEMBL IDs to gene symbols. In cases where an ENSEMBL ID did not map to a gene symbol, the ENSEMBL ID was maintained as the gene identifier. Then all gene symbols were harmonized using the limma package in R. For proteins, we either used gene mappings provided in meta-data (UKB proteomics) or ENSEMBL IDs provided by the original source (all brain proteomics). Then we followed the same procedure to harmonize the mapped genes.

**Gene Ontology Enrichment Analyses of Prioritized Drug Targets**

We generated a list of prioritized potential drug targets for each GWAS trait by combining evidence across all molecular trait analyses. With these lists of associated molecular traits, we performed gene ontology enrichment analyses for each GWAS trait separately using the R package clusterProfiler.

### Supplementary Figures

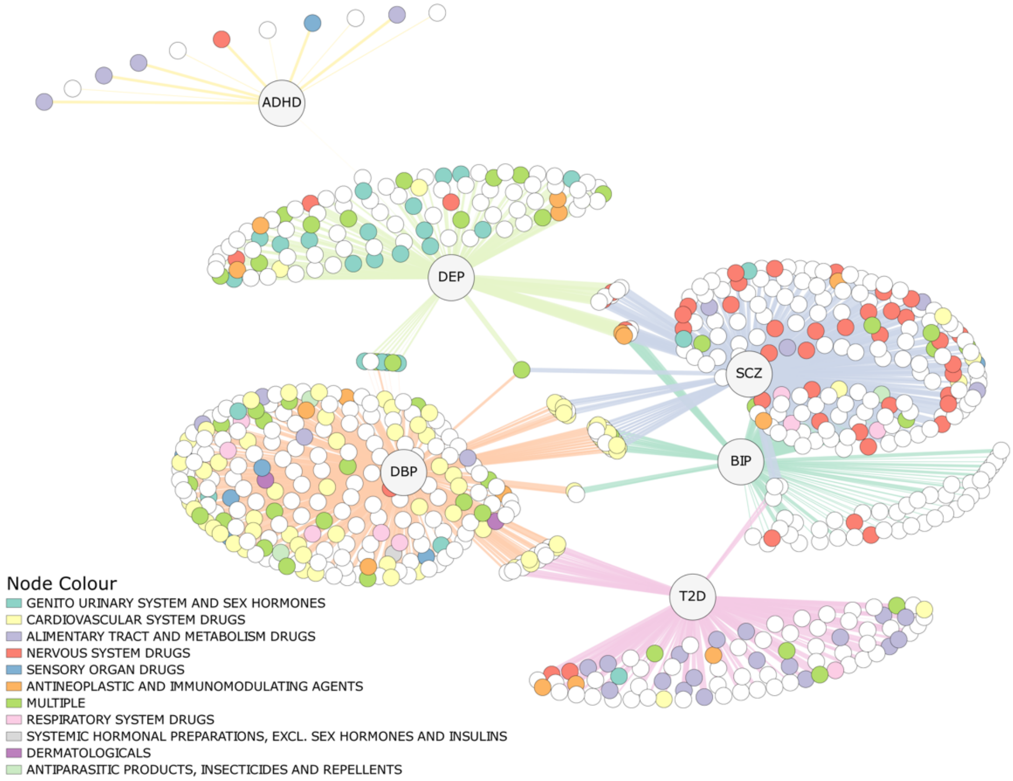


**Supplementary Figure 1. Overlap of Enriched Drugs for each GWAS Traits.** A network plot depicting each of the enriched drugs (colored nodes) for each GWAS trait labelled central nodes. Each drug node is colored based on their level 1 anatomical therapeutic chemical classification. Those nodes without a color (white) represent drugs not assigned an anatomical therapeutic chemical code. The thicker the edge (line connecting nodes to disorder), the larger the fold enrichment. ADHD = Attention deficit hyperactivity disorder, BIP = Bipolar disorder, DBP = Diastolic blood pressure, DEP = Depression, SCZ = Schizophrenia, T2D = Type 2 Diabetes.

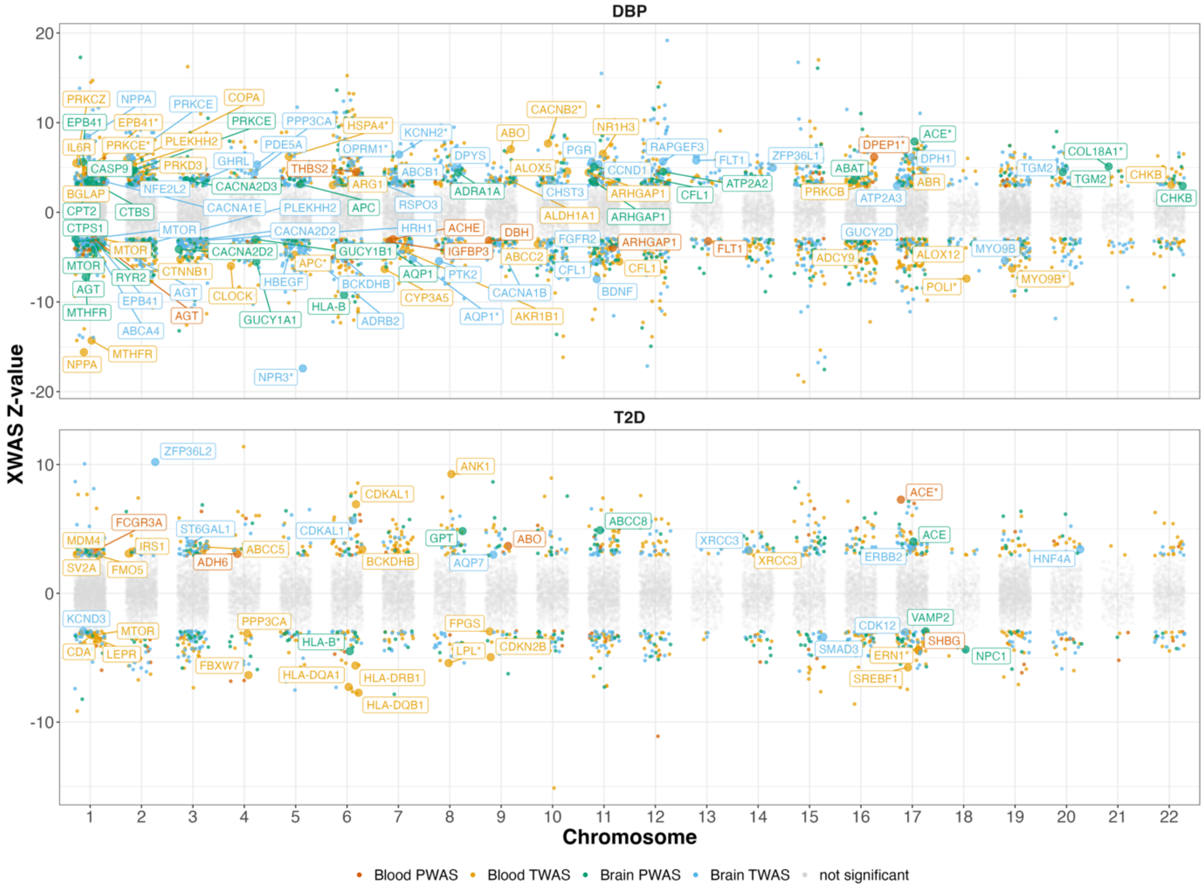


**Supplementary Figure 2. Transcriptome and Proteome Wide Association Study Results for Comparator Traits**. Significantly associated genes and proteins that are also a part of gene sets for enriched drugs are labelled. Labelled molecular traits that were also colocalized are represented with an asterisk. DBP = Diastolic blood pressure, T2D = Type 2 Diabetes, TWAS = Transcriptome wide association study, PWAS = Proteome wide association study.

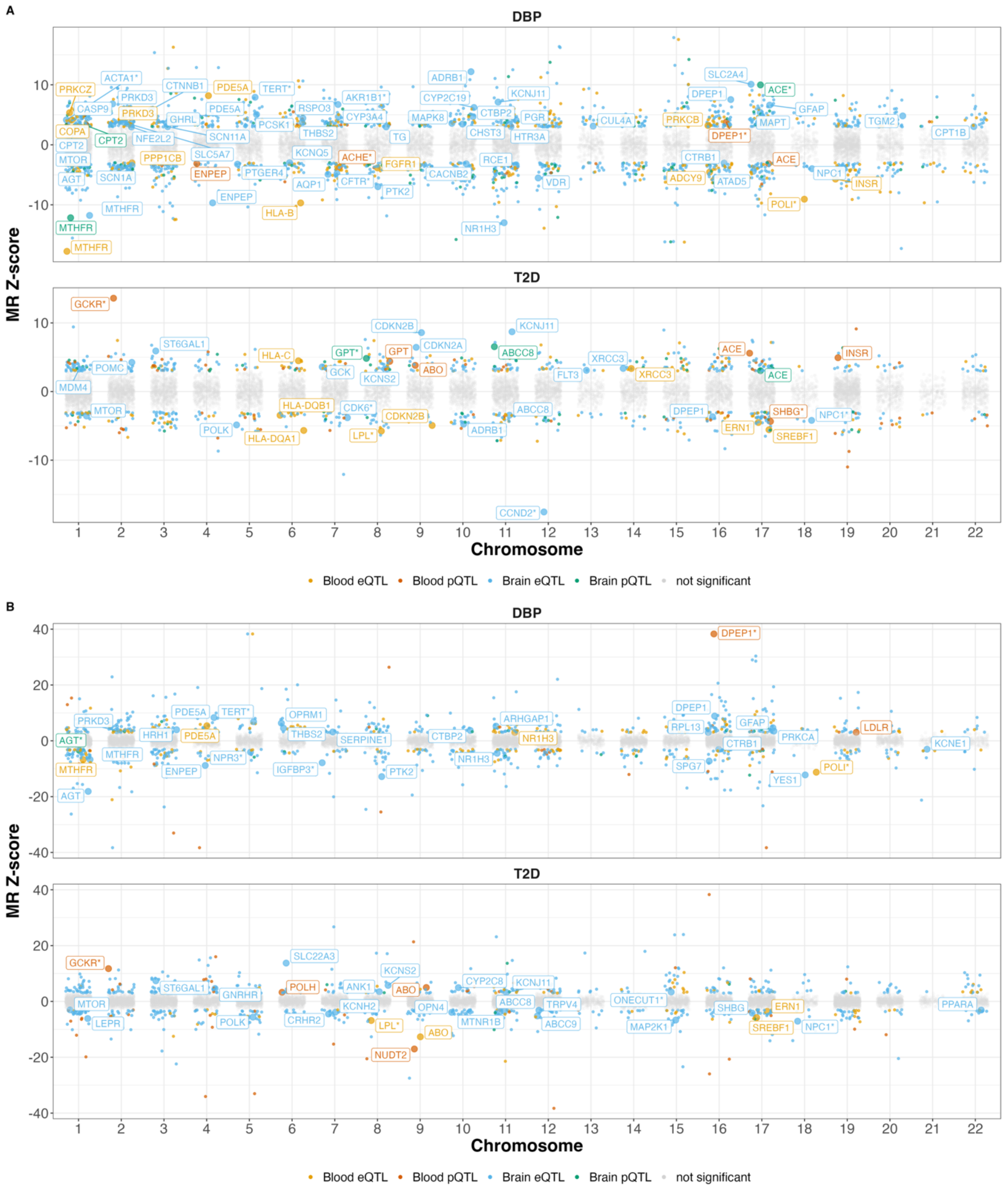


**Supplementary Figure 3. Mendelian Randomization Results for Comparator Traits**. A) shows the Mendelian randomization results in the forward direction with diastolic blood pressure (DBP) and type 2 diabetes (T2D) as the outcome traits. B) shows the Mendelian randomization results in the reverse direction with DBP and T2D as the exposure traits. Significantly associated genes and proteins that are also a part of a drug gene set enriched for DBP or T2D are labelled. Labelled genes/proteins that were also colocalized are represented with an asterisk. MR = Mendelian randomization, eQTL = expression quantitative trait loci, pQTL = protein quantitative trait loci.
